# A microplanning strategy to improve door-to-door health service delivery: The case of Seasonal Malaria Chemoprevention in Sub-Saharan African villages

**DOI:** 10.1101/2020.09.22.20199596

**Authors:** André Lin Ouédraogo, Julie Zhang, Halidou Tinto, Innocent Valéa, Edward A. Wenger

## Abstract

**Background:** Malaria incidence has plateaued in Sub-Saharan Africa despite Seasonal Malaria Chemoprevention’s introduction. Community health workers use a door-to-door delivery strategy to treat children with SMC drugs, but for SMC to be as effective as in clinical trials, coverage must be high over successive seasons.

**Methods:** We developed and used a microplanning model that ‘utilizes raster to estimate population size, generates optimal households visit itinerary, and quantifies SMC coverage based on CHWs’ time investment for treatment and walking. CHWs’ performance under current SMC deployment mode was assessed using CHWs’ tracking data and compared to microplanning in villages with varying demographics and geographies.

**Results:** Estimates showed that microplanning significantly reduces CHWs’ walking distance by 25%, increases the number of visited households by 36% (p < 0.001) and increases SMC coverage by 21% from 37.3% under current SMC deployment mode up to 58.3% under microplanning (p < 0.001). Optimal visit itinerary alone increased SMC coverage up to 100% in small villages whereas in larger or hard-to-reach villages, filling the gap additionally needed an optimization of the CHW ratio.

**Conclusion:** We estimate that for a pair of CHWs, the daily optimal number of visited children (assuming 8.5mn of treatment duration per child) and walking distance should not exceed 45 and 5km respectively. Our work contributes to extend SMC coverage by 21-63% and may have broader applicability for other community health programs.

## Background

Malaria remains the foremost health challenge in Sub-Saharan Africa [1]. Recent data showed that globally, progress in reducing malaria burden has stalled, especially in high-burden countries [1] urging the World Health Organization (WHO) to launch the country-led high burden to high impact (HBHI) approach. The goal of the HBHI is to bring the 11 highest burden countries, 10 in Sub-Saharan Africa plus India, back on track to achieve WHO Global Technical Strategy’s milestone which aims to reduce incidence by at least 75% by 2025 [2]; but that is unlikely to succeed unless key burden reduction strategies such as seasonal malaria chemoprevention (SMC) are revisited to maximize impacts.

Clinical trials reported that SMC prevent approximately 80% of malaria episodes among treated children [3-5]. At the global scale, modeling studies suggest that millions of malaria cases and thousands of deaths could be averted if SMC delivery on the ground was successful [6]. With regard to SMC deployment, studies comparing fixed-location versus door-to-door suggest the latter as most effective [7-9]. After its recommendation by WHO in 2012, SMC was deployed in 2014, mostly in countries across the Sahel and Sahel sub-region where more than 60% of clinical malaria concentrate within 3-to-4 consecutive months. Nevertheless, recent data showed that SMC in its programmatic phase is failing as progress in reducing incidence has plateaued to date despite its introduction [1].

One likely reason SMC is not properly working under real-world conditions is to be associated to its poor delivery in the community. In Mali, the average of SMC coverage in 2016 as reported from seven surveys was 53% [10]. In Burkina Faso, post-campaign coverage estimates using SMC cards and parents’ statements showed that only a small fraction of children (32%) received all SMC doses over four consecutive rounds [11]. However, for SMC to reach the desired cases reduction, we must see high coverages above 90-95% over successive seasons [6, 12]. Multiple logistic constraints and shortcomings in SMC deployment including CHW ratio per capita [11], excess time loss during treatment [5], and importantly missed households or settlements as reported during Polio vaccination [13-15] likely contribute to lower SMC coverage.

Here we develop a microplanning model to predict CHW’s performance during door-to-door health service delivery and review opportunities to optimize and standardize SMC deployment that may contribute leverage its potential and accelerate malaria burden reduction toward 2025. The model utilizes population raster data on demographics (family sizes, household geolocations) to assess treatment duration and door-to-door travel times allowing for quantification of the CHW’s time investment that is convertible to actual SMC coverage and unmet needs. We then use the model to assess SMC coverage under its current deployment mode and to predict microplanning achievements in African remote villages.

## Methods

### Ethics Statement

Data presented here were from publicly available sources and did not require ethical clearance.

### Study site

Burkina Faso reported approximately 12 million of malaria cases and 4,000 deaths in 2018 [1]. Malaria transmission is intense and seasonal [16, 17] and despite SMC, clinical malaria remains on the rise [1]. The health and demographic surveillance site (HDSS) of Nanoro in the centre west provides rich household survey data suitable for microplanning studies [18]. Three villages (Soaw, Rakolo, Mogdin) with different characteristics were selected to test the potential of microplanning in optimizing SMC deployment. Census data were obtained from the HDSS and incidence data from national DHIS2 platforms.

### Microplanning model

To improve SMC door-to-door delivery, we used a salesman algorithm-based accessibility model (Figure 1) to determine the optimal itinerary for CHWs to efficiently visit all households in each village. The model computes the shortest door-to-door visit itinerary using global positioning system (GPS) information of households and outputs travel distance and travel time as well as treatment duration. Households’ GPS coordinates and family sizes were extracted from the population raster of each village. Population raster of Burkina Faso as provided by CIESIN and the Connectivity Lab at Facebook encapsulates values on number of individuals inside raster’s pixels that can be processed to extract family sizes [19]. Fraction of under 5 children per household was subsequently derived from related-family size assuming that under 5 children make up approximately 18% of the total population [20].

**Figure 1:**
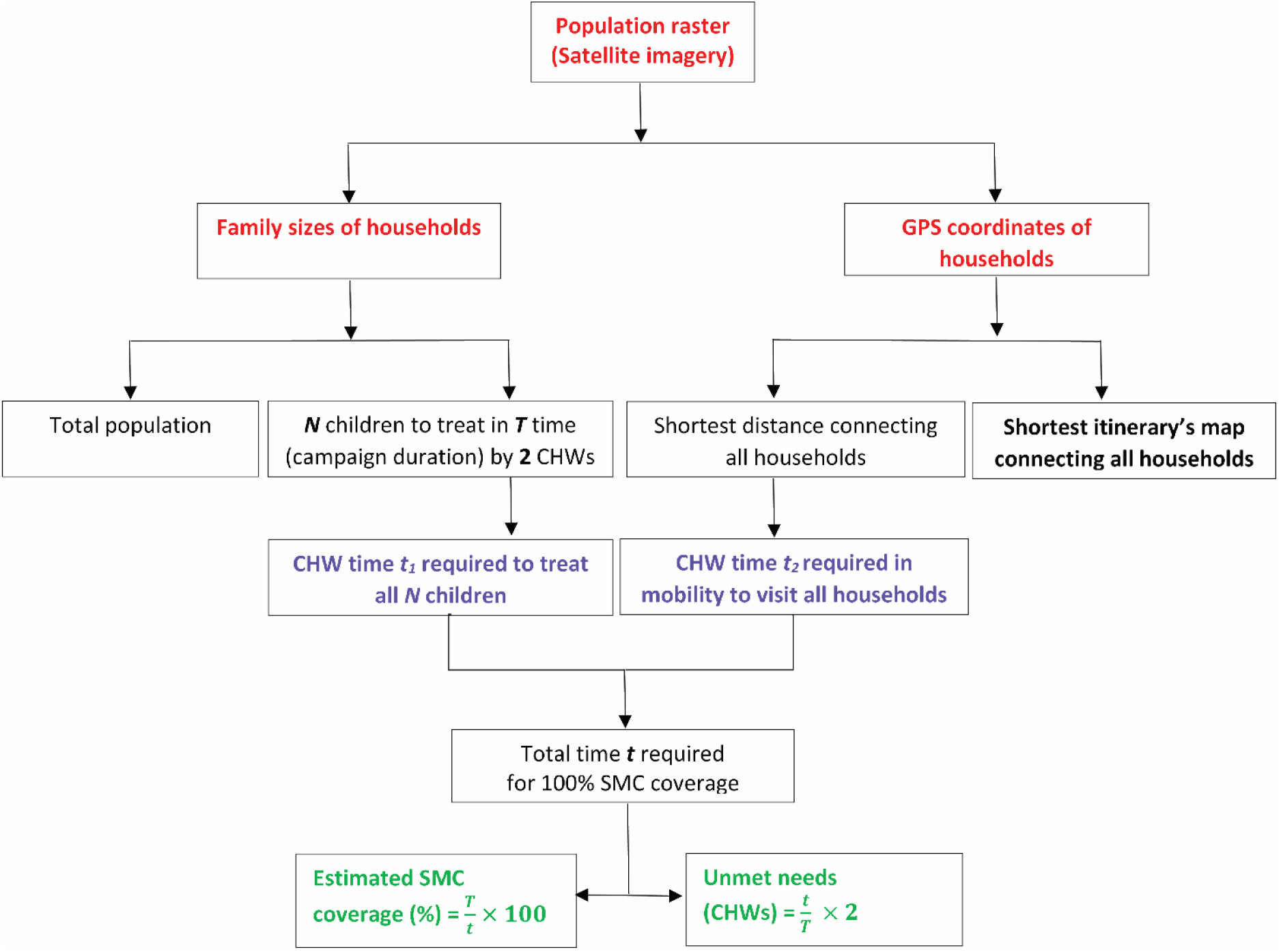
Microplanning model design.

#### Generating household visit itinerary using the salesman algorithm

We describe the traveling salesman problem (TSP) as a graph theory problem. Each household is thought of as a vertex and each vertex is connected by an edge, so our graphs is *G =* (*V, E*), where *V* is the set of vertices, and *E* is the set of edges. Each edge has an associated cost *c*_*ij*_, which is the distance between the two households. Our goal is to find the shortest path from any starting vertex that passes through all other vertices without repeating. Unlike the classic TSP, we do not return to the starting household. Instead, we use the Held-Karp algorithm, a dynamic programming solution [21]. The idea is to compute optimal sub-paths. We compute table entries C(S, i, j) for each subset S ⊂ V, and i, j ∈ S, defined to be the length of the shortest path from vertex *i* to vertex *j* visiting each vertex in *S* exactly once (and no node outside of *S*). The algorithm computes *C*(*S, i, j*) for increasing number of vertices in set *S*, up to *N*, the total number of vertices.

1. Let C({i, j}, i, j*) =* c_ij_ for all i ≠ j.
2. For *K = 3* to *N*, do
  a. For all sets S ⊂ V with *K* vertices, compute 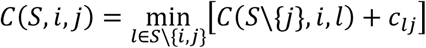
  b. Return the optimal cost 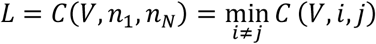

We now can recover the path as follows: *n*_1_, *n*_*N*_ are our starting and ending vertices respectively. Vertex n_N−1_ is the unique vertex satisfying

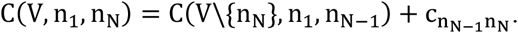

If we have computed n_N−1_, …, n_j+1_, then vertex *n*_*j*_ is the unique vertex satisfying

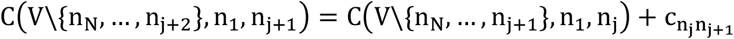

We now have the whole path (*n*_1_, …, *n*_*N*_) along with optimal distance *L = C*(*V, n*_1_, *n*_*N*_). With the path linking household via GPS coordinates, we could estimate walking distance using Euclidian distance formula.

#### Subdividing hard-to-reach areas using k-means clustering

For hard-to-reach villages with accessibility constraints (e.g. rivers), the model first clusters households using the constrained K-Means algorithm before determining optimal itinerary and unmet needs for each cluster [22].

We give the mathematical formulas for the constrained K-Means problem. Let the dataset be *D =* {*x*_1_, …, *x*_*m*_}, where *x*_*i*_ ∈ *R*^*n*^. Let 1 ≤ *K* ≤ *m* be the number of clusters. We want to find cluster centers *C*_1_,.., *C*_*K*_ such that the distance between each point *x*_*i*_ and the nearest cluster center *C*_*h*_ is minimized under the condition that cluster number *h* must contain at least *τ*_*h*_ data points, where 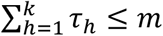. If *τ* > 0, this forces clusters to be non-empty, and we can also choose *τ*_*h*_ such that all clusters have relatively the same number of data points. We let T_i, h_ ∈ {0, 1} denote the “selection variables” that indicate whether *x*_*i*_ belongs to cluster number *h*. The constrained K-Means problem is as follows.

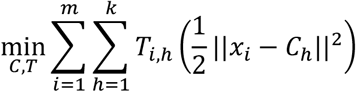

We can solve this iteratively. At iteration *t*, let C_1, t_, …, C_k, t_ be the cluster centers. We compute the cluster centers C_1, t+1_, …, C_k, t+1_ at iteration t + 1 in 2 steps.

1. Cluster Assignment: Let 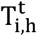 be a solution to the following linear program with C_h, t_ fixed.

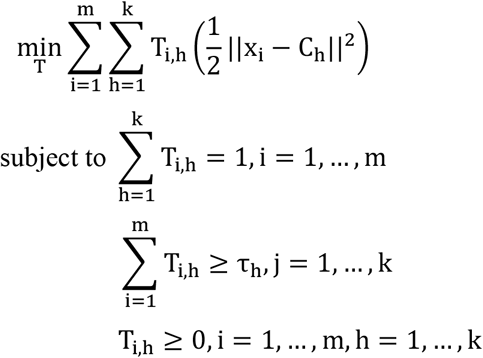
2. Update C_h, t+1_ as follows. If 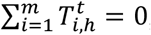, then no update is made: *C*_*h,t*+1_ = *C*_*h,t*_. If 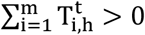, then

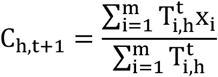

We terminate when C_h, t+1_ *=* C_h, t_ for all *h*. The constraints in the linear program in the cluster assignment step is equivalent to a Minimum Cost Flow (MCF) problem, a linear network optimization problem.

### SMC performance under current standard deployment

Current standard SMC deployment refers to as a door-to-door delivery performed by a CHW whose geographical orientation and time management is solely based on the CHW’s own perception. SMC coverage was defined as follows: 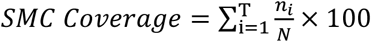 where T is campaign duration in days, *n* is number of treated and N the total number of children. Based on personal communications and on reports, we estimated at 12.5mn the average treatment duration per child ranging under 15mn to above 30mn in 63% to 22% of occasions respectively [23]. During a SMC campaign, CHWs service packages were loaded with GPS devices and unknowingly provided GPS-tracking itineraries in Rakolo. Walking distances were estimated using visited household coordinates and converted to travel times assuming a 20mn walk per km in the wet season [24, 25].

### SMC performance under microplanning

To predict SMC coverage under microplanning, the model assumes an initial number of 2 CHWs, daily working time of 8 hours, and 4 days of campaign duration. Walking distances in optimized visit itineraries were converted to travel times [24, 25]. Based on current CHWs experiences, we assumed random draws of treatment duration for 1, 2 or 3 children per household as follows t ∼ U (10, 15); t ∼ U (15, 20) or t ∼ U (20, 25) respectively. Assuming a household of 3 children we estimated on average 8.5mn (25mn/3) per child as best optimal treatment duration. Predicted SMC coverage was computed as follows:

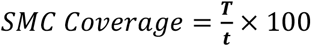 where T is campaign duration in days and t is total of treatment and travel times in days (Figure 1).

Comparison analyses of proportions of treated children and visited households between current SMC deployment and microplanning were based on Chi^2^ test.

### Unmet needs for SMC performance maximization

To maximize SMC performance, unmet needs were estimated by converting maximum time invested to reach 100% of SMC coverage into number of CHWs needed:

Unmet needs 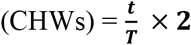 where T is campaign duration in days and t is total time invested for treatment and travel (Figure 1). We chose to express unmet needs as supplementary CHWs instead of supplementary campaign days to reduce workforce burden (fatigue).

## Results

### Predicting population sizes using raster

Three villages with varying characteristics were selected to assess CHWs’ performances and unmet needs under current SMC deployment mode and two microplanning scenarios (A and B). In Rakalo, households seem evenly spaced suggesting a uniform dispersion of the population (Figure 2). Contrarily to Rakolo, households in Mogdin are unevenly spaced leading to a random dispersion of the population with longer distances to connect households (Figure 2). In Soaw, households are distributed across water streams leading to clumped dispersion of the population suggesting that hard-to-reach scenarios should be accounted for while microplanning SMC delivery. We thus subdivided Soaw into three groups (A, B, C, Figure 2) using k-means clustering. Predicted under 5 population sizes in Rakolo, Mogdin and Soaw using raster were not significantly different (146, 324, 945) compared to census data (146, 331, 998 respectively; Figure 3A).

**Figure 2:**
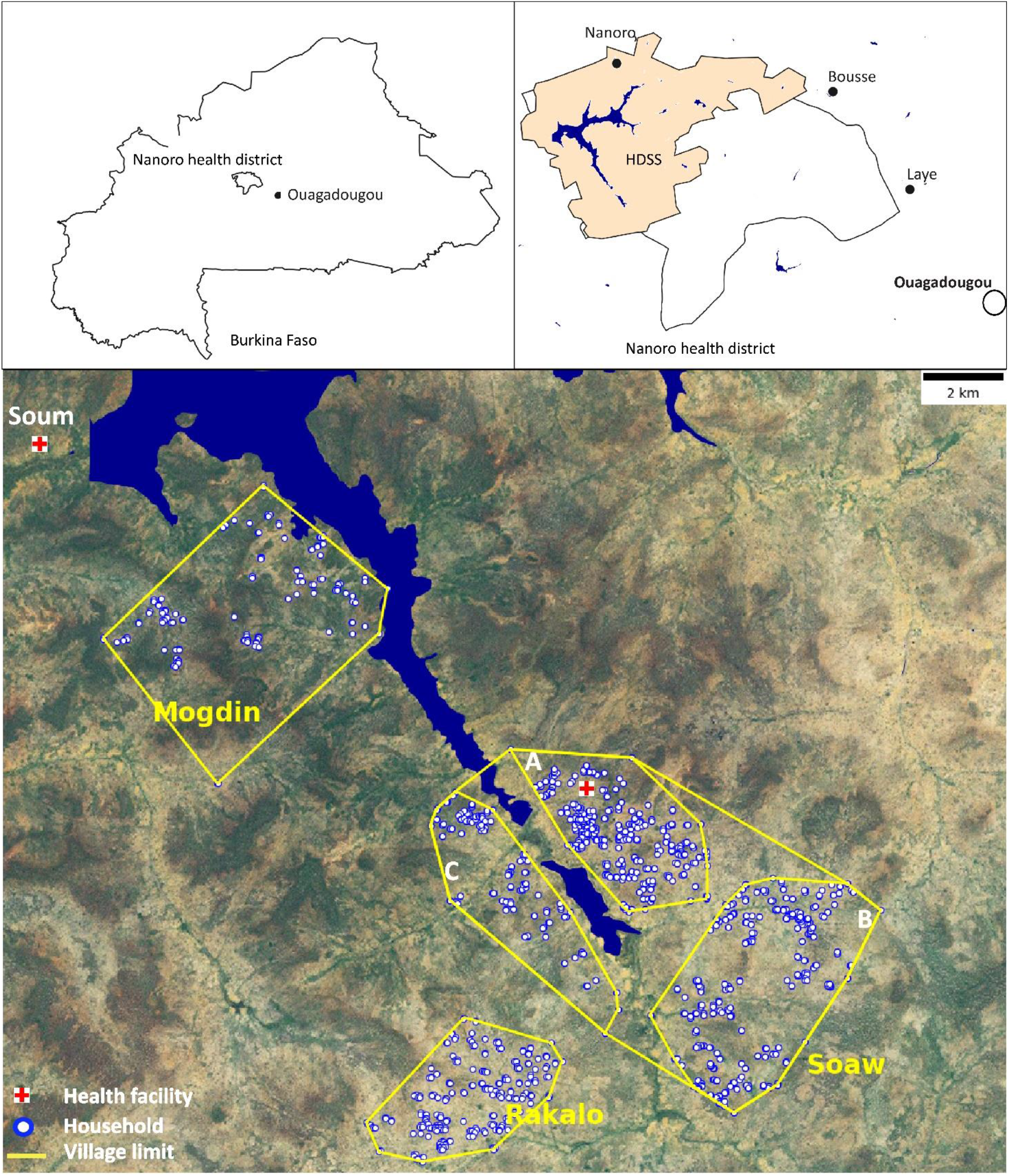
Study villages.

**Figure 3:**
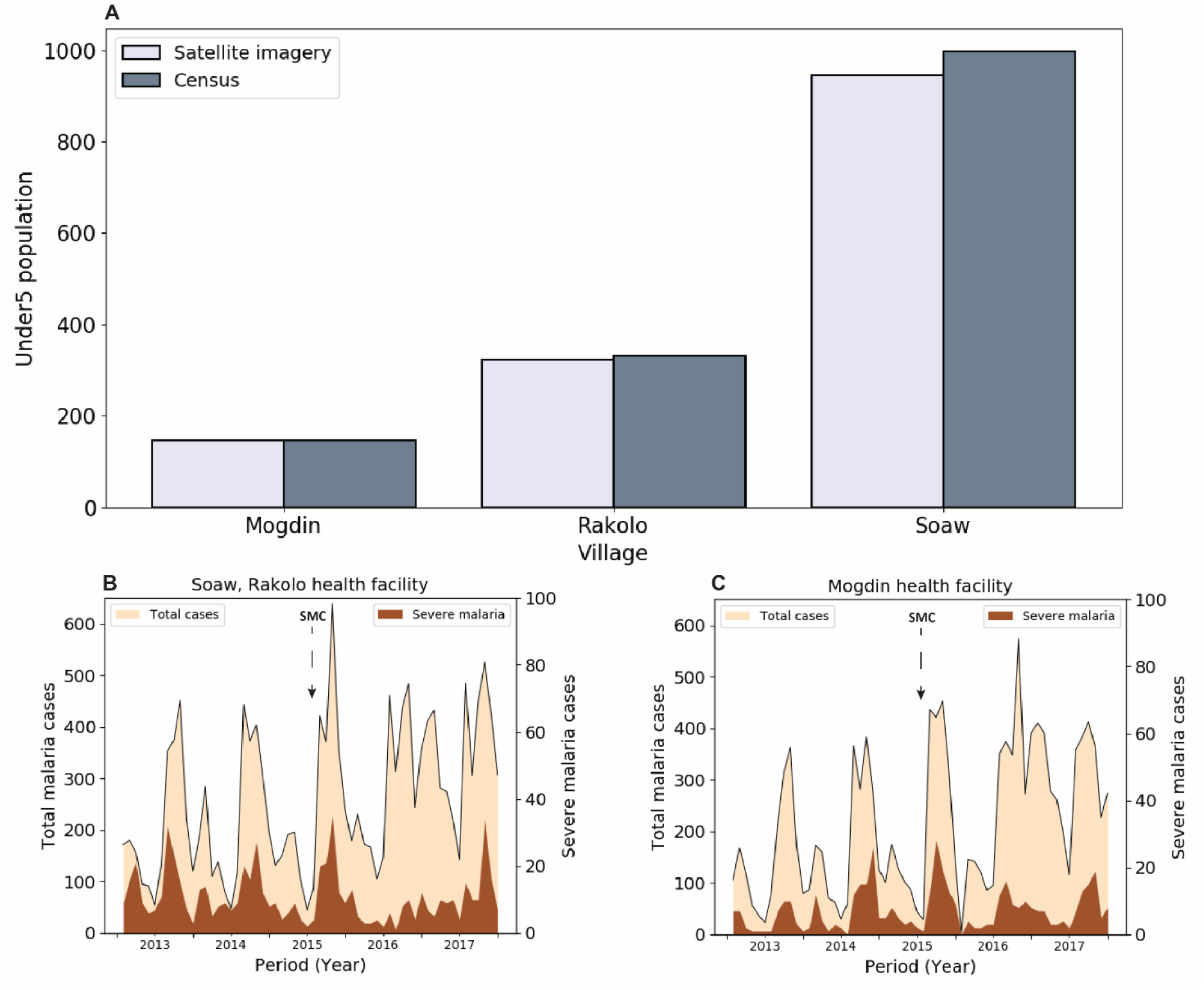
Population estimates and malaria burden in study villages.

### Malaria incidence not impacted by current SMC deployment

Routine incidence data of Soum’s primary health facility catchment area was assumed from at least both villages of Soum and Bogdin (Figure 1). Data reported by Soaw primary health facility were assumed from at least both villages of Soaw and Rakolo. Note that routine data outside catchment area are likely reported by our health facilities and conversely but it is difficult to estimate a differential incidence data for each village. Temporal malaria incidence increased or stalled in all three villages as from 2013 to 2017 suggesting that SMC deployment has likely not been effective to date (Figure 3B, 3C). Similar incidence trends were observed from other health facilities supporting that SMC is likely not impactful in the entire region (Supplementary Figure 1).

### Impact of microplanning on SMC coverage

CHWs’ performance including number of visited households, number of treated children, walking distance and ultimately SMC coverage were estimated under current SMC deployment mode and compared to microplanning modes (Figure 4A, B). Performances were predicted using raster-based population estimates. Two types of microplanning (A, B), both assuming an optimal treatment duration of 8.5mn per child, were introduced to improve current SMC deployment. A microplanning A consisted of providing the CHW with a household visit itinerary plan which helps visualize the extent of the catchment area on a map with the shortest itinerary to visit all households. When microplanning A was predicted ineffective to cover 100% of households within the given campaign duration of 4 days, a microplanning B was introduced which consisted of optimizing (adjusting) the number of CHWs to timely cover all households and treatments.

**Figure 4:**
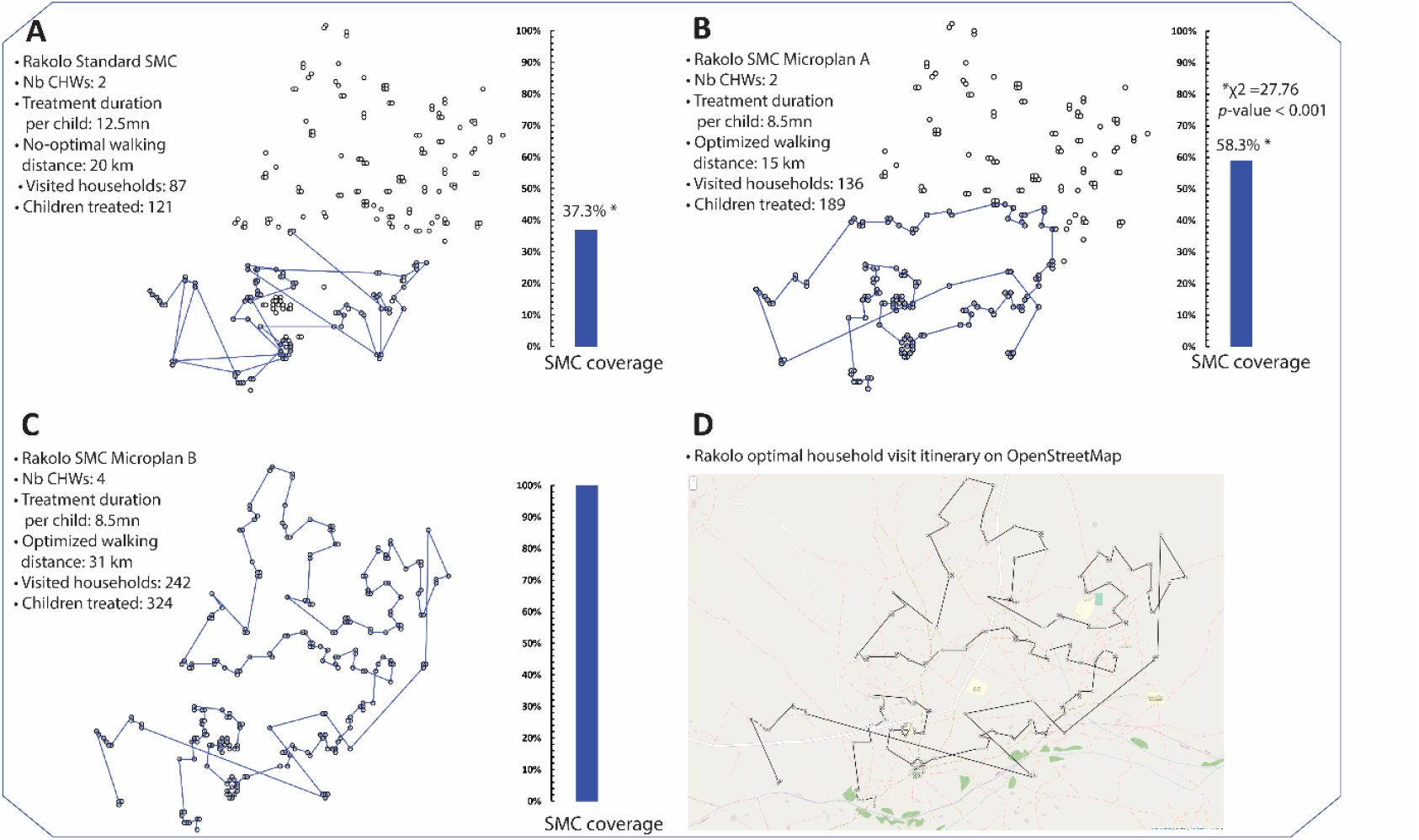
Performance of current SMC deployment mode (A) compared to microplanning (B, C) in Rakolo. Microplan A = Visits itinerary and time invested in treatment are optimized; Microplan B = Visits itinerary, time invested in treatment and number of CHWs are optimized.

Based on CHWs’ tracking data we estimated 37% of SMC coverage (Figure 4A) in Rakolo which is in line with previous studies [11]. A total of 136 households were visited by CHWs when a visit itinerary plan was introduced (microplanning A, Figure 4B) which significantly reduced CHWs’ walking distance by 25% (20km to 15km). This represents a significant increase by 36% (*χ2* = 19.15, p < 0.001) in the number of visited households as compared to the CHWs’ tracking data under current SMC deployment mode (87 households visited, Figure 4A). The equivalent in SMC coverage was a significant increase by 21% (*χ2* =27.76, p < 0.001, Figure 4) as the number of children treated increased from 37.3% (121/324) under current SMC deployment mode up to 58.3% (189/324) following the introduction of optimal visit itinerary plan. Maximization of SMC coverage to 100% within 4 days window required 2 additional CHWs as unmet need in the roll out of microplanning B to extend the reach of SMC treatment (Figure 4C). The practicality of using a model-optimized visit itinerary plan for SMC deployment in Rakolo is illustrated in Figure 4D.

CHWs’ tracking data were not available for Mogdin and Soaw and therefore SMC coverage under current SMC deployment was not assessed. Microplanning A was systematically applied to Mogdin and Soaw and replaced by a microplanning B if 100% of SMC coverage was not reached under the former.

In Soaw, many children in both clusters A (277 children) and B (235 children) were missed under microplanning A (Table 1). Unmet needs were estimated to be 2.38 and 2.12 additional CHWs required for cluster A and B respectively (Figure 5A, 5B). In cluster C, an optimal visit itinerary alone (microplanning A) was enough to 2 CHWs to complete treatment of 146 children alongside 20km of walking (Figure 5C). Similarly, the use of an optimal visit itinerary alone in Mogdin was enough to cover all 146 eligible children in less than 4 days (Figure 5D). The practicality of using a model-optimized visit itinerary plan for SMC deployment in Soaw is illustrated in Supplementary Figure 2.

**Table 1.** Performances of SMC under various deployment modes within 4 days of SMC campaign

| Village | SMC Deployment mode | CHWs needed<br>(Visited households) | Children treated<br>(Treatment time) | Walking distance in<br>km (Travel time) | SMC coverage<br>(Treated/Total) | P-value |
| --- | --- | --- | --- | --- | --- | --- |
| Rakolo | Current (Ref.) | 2 (87) | 121 (3.15 days) | 20 (0.83 days) | 37.3% (121/324) |  |
|  | Microplan A | 2 (136) | 189 (3.35 days) | 15 (0.63 days) | 58.3% (189/324) | <0.001 |
|  | Microplan B | 3.5 (242) | 324 (5.74 days) | 31 (1.29 days) | 100% (324/324) | <0.001 |
| Mogdin | Microplan A | 1.87 (126) | 146 (2.58 days) | 28 (1.16 days) | 100% (146/146) |  |
| Soaw A | Microplan A | 2 (152) | 195 (3.45) | 13.2 (0.55 days) | 45.6% (195/429) |  |
|  | Microplan B | 4.38 (323) | 429 (7.6 days) | 36 (1.16 days) | 100% (429/429) |  |
| Soaw B | Microplan A | 2 (135) | 179 (3.16) | 20.16 (0.84 days) | 48.5% (179/370) |  |
|  | Microplan B | 4.12 (278) | 370 (6.55 days) | 41 (1.70 days) | 100% (370/370) |  |
| Soaw C | Microplan A | 1.7 (118) | 146 (2.58 days) | 20 (0.83 days) | 100% (146/146) |  |
| Average |  | 2 (135) | 178 (3.238 day) | 19.03 (0.762 day) |  |  |

Current = Current SMC deployment mode on the ground, Microplan A = Visits itinerary and time invested in treatment are optimized; Microplan B = Visits itinerary, time invested in treatment and number of CHWs are optimized

**Figure 5:**
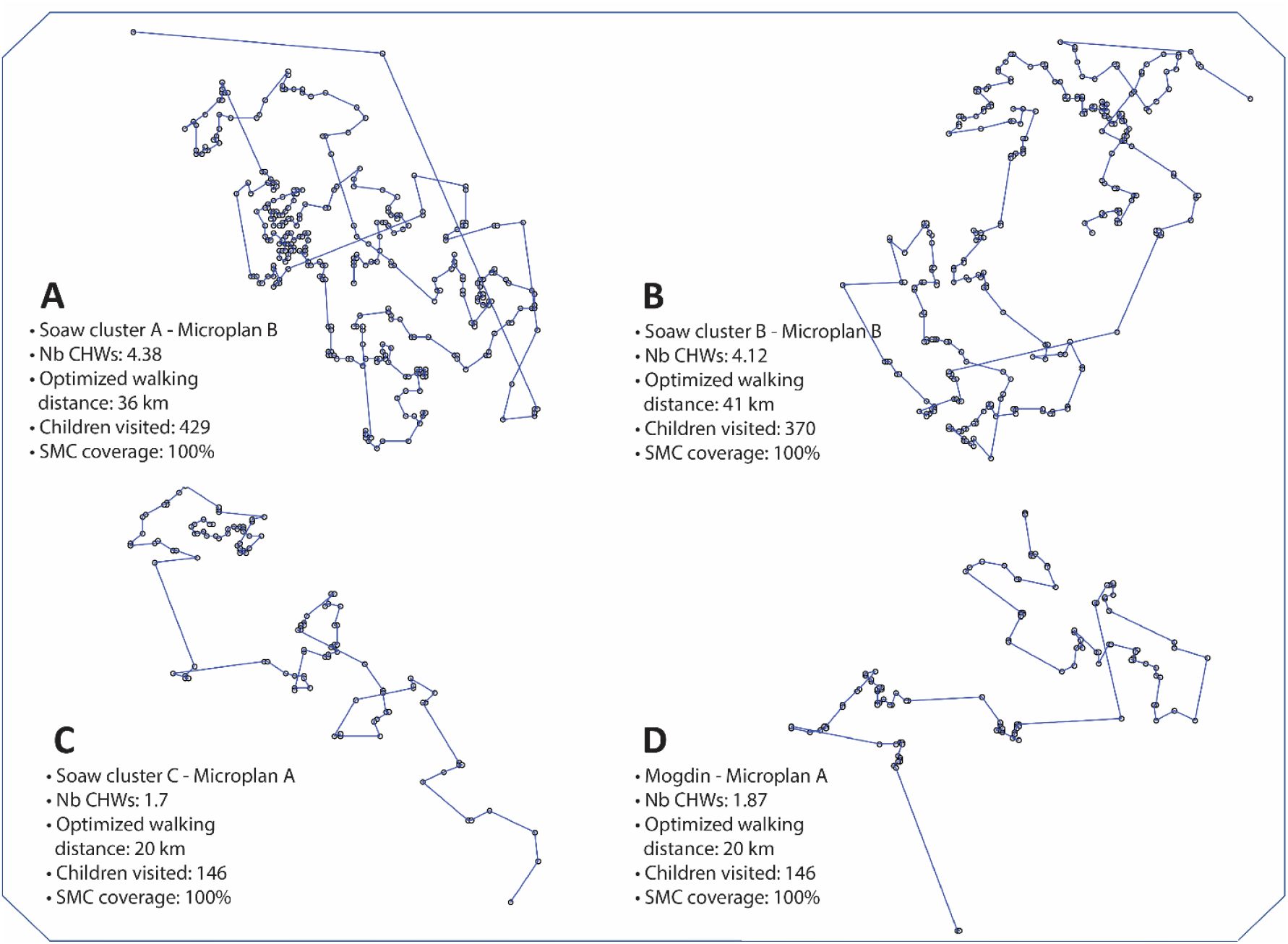
Performance of SMC microplanning in Soaw (A, B, C) and Mogdin (D). Microplan A = Visits itinerary and time invested in treatment are optimized; Microplan B = Visits itinerary, time invested in treatment and number of CHWs are optimized.

On average we estimated that for a pair of CHWs, the daily optimal number of children for treatment and walking distance should not exceed 45 and 5km respectively.

## Discussion

The present work identified opportunities to extend the reach of SMC treatment in rural African villages. Key inputs to SMC deployment including household visit itinerary, treatment duration and CHW ratio per capita were revisited under microplanning strategies. On average we estimated that for a pair of CHWs, the daily optimal number of children for treatment and walking distance should not exceed 45 children and 5km respectively over 4 days of campaign. We showed that optimal household visit itinerary reduces CHWs’ walking distance by 25%, increases the number of visited households by 36% and increases SMC coverage by 21% (37% under current standard mode up to 58% under microplanning). SMC coverage could be maximized to 100% only when the number of CHWs was adjusted to proportionate to the number of eligible children.

### Microplanning household visit itinerary

Our estimate of 37% of SMC coverage based on CHWs’ door-to-door tracking data is in line with previous findings [11] and possibly reflects current poor performance of SMC in the area. For millions of children in Sub-Saharan Africa, the door-to-door service delivery is a relief and often seen by stakeholders as an equity-focused strategy to leverage. In 2014, door-to-door delivery was deployed to maximize SMC coverage across the Sahel region, particularly to outreach those that are furthest from health facilities. But as with other interventions such as vaccination campaigns, the most vulnerable remain those that are remote and likely to be missed due to poor logistics or lack of microplanning. Our model estimates 21 - 63% increase in SMC coverage following the introduction of microplanning as 36% of households, previously missed by CHWs under current SMC deployment mode were recovered by microplanning. Using tracking systems and GIS-based microplanning, polio eradication teams were able to recover up to 38% of missed settlements from 8 Nigerian’s states during routine vaccine campaigns in 2013 [13] and an outreach of 31-43% (140 out of 322 – 441 settlements) during supplement immunization campaigns in Kano state [15]. Missing households or entire settlements during door-to-door service delivery is not uncommon in rural areas in Sub Saharan Africa where most households are not connected with roads. Without maps of catchment areas and having to crisscross winding trails and bushes, CHWs’ teams will not only walk longer distances but likely will miss households. A recent report highlights that the lack of planned itineraries and similarities between households, compounds contribute to confuse CHWs in their ability to identify the next household to be visited [14].

To be effective on the ground, SMC campaigns need to approach the excellence in delivery of clinical trials, but it must be practical and cost-effective. In clinical trials, households and participants are pre-identified and health workers trained to access them using rough sketch maps [26]. During campaigns, households are likely to be missed if biases in defining catchment areas are initially introduced in sketched paper maps or if such paper maps are not used that all [13, 15, 27].

The present work is on track to reproduce clinical trial delivery performance by focusing on tree basic needs for CHWs: go beyond rough sketching of settlements to use better microplanning of household visit itinerary, adjust CHW ratio based on population size and standardize treatment duration. Overall, our microplanning reduces treatment duration by 32% (12.5mn to 8.5mn per child) and can be standardized for better SMC performance. In Sub-Saharan Africa, census data are more often out of date or imprecise at sub-national levels while settlements identification [28-31] are not guaranteed to allow robust SMC deployment. Such biases are generally not accounted for during monitoring and evaluation efforts and often result in misleading SMC coverage estimates. The present work has the advantage to remotely assess family sizes [32] and adjust CHW ratio prior to SMC deployment. The work presents advantages of computing CHWs’ optimized travel itineraries and combining them with local accessibility features and geographies to generate printable accessibility maps or to incorporate such maps into mobile applications to be used by CHWs and supervisors.

### Limitations

Our work presents some limitations. We estimated population sizes of 2013 using a 2015 raster which resulted in less uncertainties. For further estimates, updated raster will be essential to capture population growth. The model optimizes SMC deployment on Day 1 only assuming that 2nd and 3rd SMC doses are well carried out by mothers on Day 2, 3. CHWs’ tracking data under current SMC deployment mode were only available for one village (Rakolo) while more data are needed to improve model robustness. To predict SMC coverage, we did not account for fractions of kids vomiting, refusals or those not found at home (travelers or absence). Such data will be helpful in improving predictions of SMC coverage. However, such data could be accounted for as they become available. Other challenges remain including reaching out to pastoralist and displaced populations due to conflicts. Finally, our salesman algorithm assumes catchment areas are free of physical barriers to generate free walking itineraries and therefore is currently not applicable to urban areas with excessive clustering. As next steps, we plan to include road - network processing to overcome such challenges in urban areas.

### Strengths, implications for other health programs, policy and practicality

The chronic underfunding of the health care system in low- and middle-income countries (LMCI) [33], has led to significant disparities in access to care across different settings. In Burkina Faso for instance the ratio of health infrastructure and of clinic visits per capita is estimated to be 1.03 (range 0.11 – 2.03) per 10,000 and 0.6 (range 0.08-1.01) per person per year respectively. Most deprived populations live in rural and remote settings [34]. Microplanning using satellite imagery and other geographic information systems [13, 15] are emerging strategies in door-to-door delivery and likely has recently contributed to wild polio eradication from Africa [35]. In the time of COVID-19, and to maximize the distribution of Insecticide Treated Net in COVID-19 affected countries, stakeholders for malaria prevention recently called out the need of microplanning strategies using topographic and route mapping [36] suggesting that our work may be helful. As CHWs have emerged as critical human resources to health systems in LMCI, we believe that the present work might be an opportunity to assist and improve community health programs.

To conclude, our work shows that microplanning contributes to extend SMC coverage by 21-63% in villages of Burkina Faso and may be reproducible elsewhere using free satellite raster. To the best of our knowledge, our work is first to assess opportunities of using microplanning strategies to improve SMC deployment on the ground. While this work focuses on SMC drug delivery, the microplanning strategy behind by addressing both households visit itinerary and CHW ratio per capita may have broader applicability for many other health service delivery programs such as vaccination, family planning or nutrition.

## Data Availability

Data are available upon request.

## Authors’ contributions

ALO conceived and developed the microplanning modeling framework. JZ and EW contributed to model framework development and methods. HT and IV provided household survey data, CHWs’ tracking data and malaria incidence data. ALO run the simulations an analyzed the data. ALO wrote the original draft of the manuscript. All authors contributed intellectually and made contributions to the manuscript text.

## Acknowledgments

This publication is based on research developed by the Institute for Disease Modeling prior to its affiliation with the Bill & Melinda Gates Foundation. The findings and conclusions contained within are those of the authors and do not necessarily reflect the official positions or policies of the Bill & Melinda Gates Foundation.

We thank the Ministry of Health and the primary health facility nurses and community’s health workers of Burkina for explaining their contribution to SMC campaigns in Burkina. We thank the personnel of the National Malaria Control Program in Burkina for detailed description of SMC deployment in Burkina Faso. Thank you to Karim Derra from the Clinical Research Unit of Nanoro, Burkina Faso and Kurt Frey from the Institute for Disease Modeling for helpful discussions.

---

**Supplementary Figure 1:**
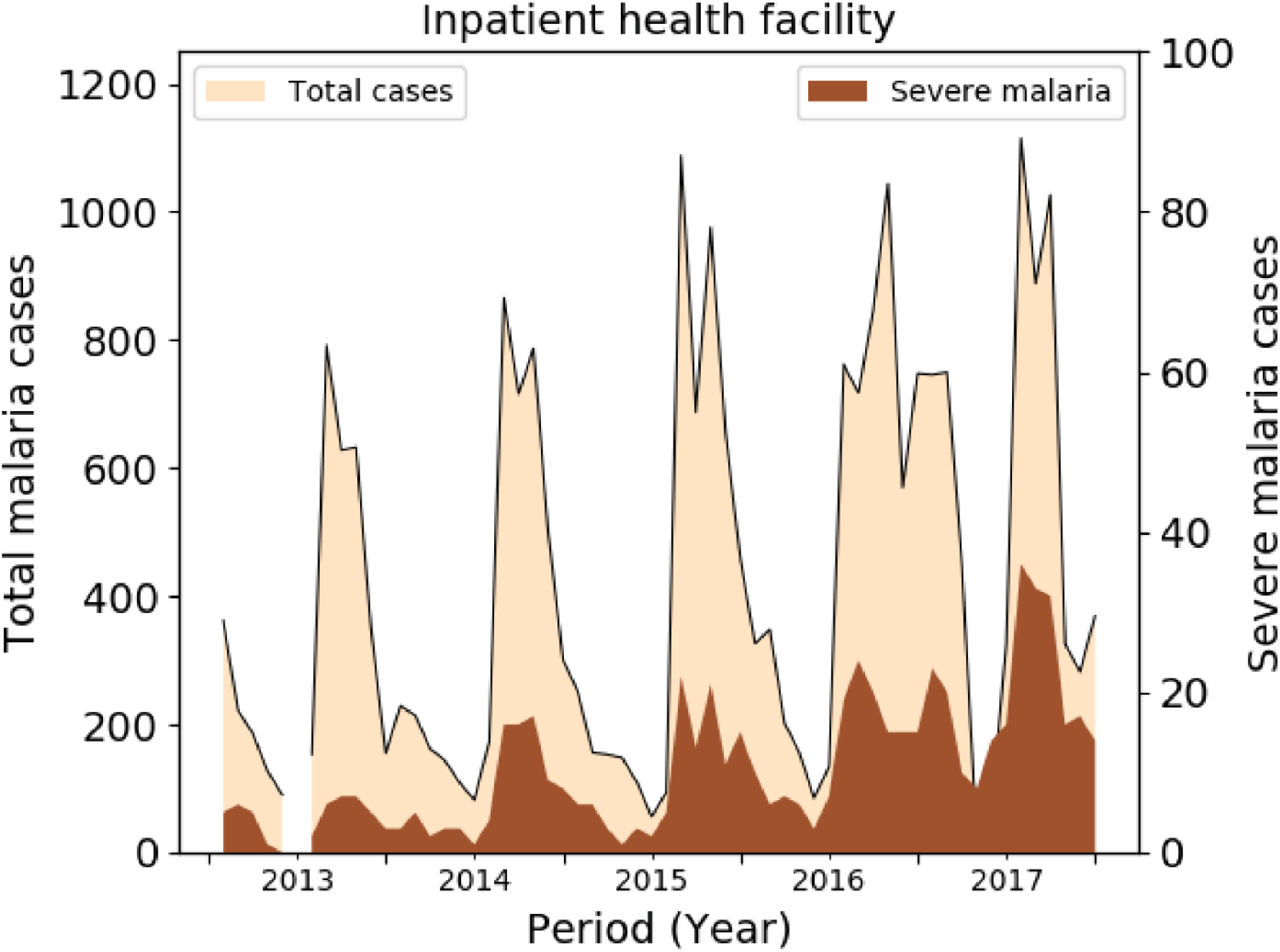
Malaria incidence as reported by the Nanoro inpatient facility.

**Supplementary Figure 2:**
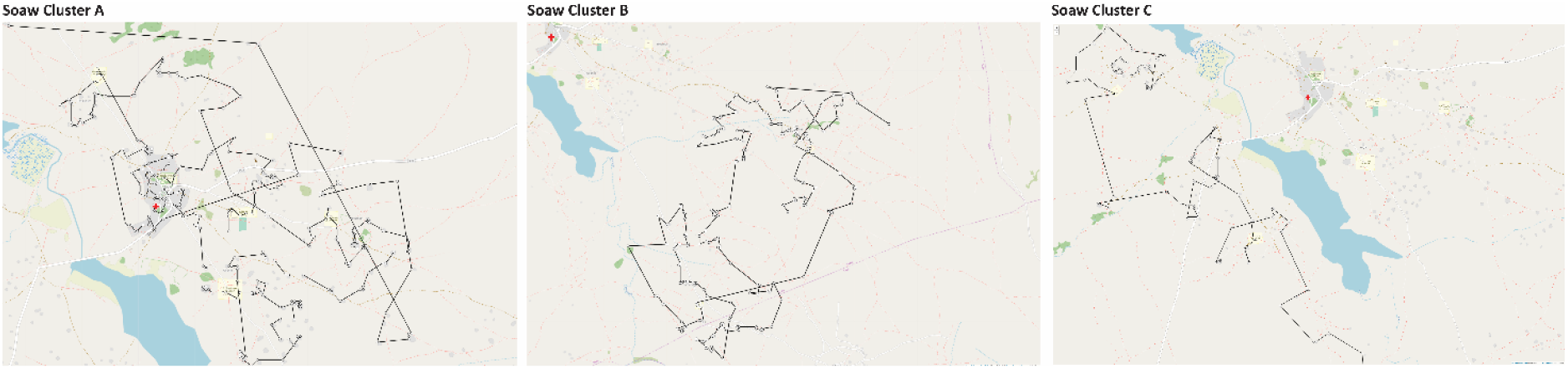
Optimized household visit itineraries over OpenStreetMap in Rakalo.

## References

1. World Health Organization. World Malaria Report. Geneva,. http://www.who.int/malaria/wmr 2019.

2. World Health Organization. Global Technical Strategy for Malaria 2016–2030. https://www.who.int/malaria/publications/atoz/9789241564991/en/ 2015.

3. Meremikwu MM, Donegan S, Sinclair D, Esu E, Oringanje C. Intermittent preventive treatment for malaria in children living in areas with seasonal transmission. Cochrane Database Syst Rev 2012:CD003756.

4. Wilson AL, Taskforce IP. A systematic review and meta-analysis of the efficacy and safety of intermittent preventive treatment of malaria in children (IPTc). PloS one 2011; 6:e16976.

5. Zongo I, Milligan P, Compaore YD, et al. Randomized Noninferiority Trial of Dihydroartemisinin-Piperaquine Compared with Sulfadoxine-Pyrimethamine plus Amodiaquine for Seasonal Malaria Chemoprevention in Burkina Faso. Antimicrobial agents and chemotherapy 2015; 59:4387–96.

6. Cairns M, Roca-Feltrer A, Garske T, et al. Estimating the potential public health impact of seasonal malaria chemoprevention in African children. Nature communications 2012; 3:881.

7. Barry A, Issiaka D, Traore T, et al. Optimal mode for delivery of seasonal malaria chemoprevention in Ouelessebougou, Mali: A cluster randomized trial. PloS one 2018; 13:e0193296.

8. Bojang KA, Akor F, Conteh L, et al. Two strategies for the delivery of IPTc in an area of seasonal malaria transmission in the Gambia: a randomised controlled trial. PLoS medicine 2011; 8:e1000409.

9. Kweku M, Webster J, Adjuik M, Abudey S, Greenwood B, Chandramohan D. Options for the delivery of intermittent preventive treatment for malaria to children: a community randomised trial. PloS one 2009; 4:e7256.

10. Diawara F, Steinhardt LC, Mahamar A, et al. Measuring the impact of seasonal malaria chemoprevention as part of routine malaria control in Kita, Mali. Malaria journal 2017; 16:325.

11. Compaore R, Yameogo MWE, Millogo T, Tougri H, Kouanda S. Evaluation of the implementation fidelity of the seasonal malaria chemoprevention intervention in Kaya health district, Burkina Faso. PloS one 2017; 12:e0187460.

12. WHO World Health Organization. Guidelines for the treatment of malaria. [http://www.who.int/malaria/docs/TreatmentGuidelines] 2006.

13. Barau I, Zubairu M, Mwanza MN, Seaman VY. Improving polio vaccination coverage in Nigeria through the use of geographic information system technology. The Journal of infectious diseases 2014; 210 Suppl 1:S102–10.

14. Malaria Consortium. Notes from a site visit to a seasonal malaria chemoprevention (SMC) program supported by Malaria Consortium in Burkina Faso, August 18-22, 2019. https://files.givewell.org/files/conversations/Malaria_Consortium_site_visit_in_Burkina_Faso_August_18_22_2019.pdf 2017.

15. Gali E, Mkanda P, Banda R, et al. Revised Household-Based Microplanning in Polio Supplemental Immunization Activities in Kano State, Nigeria. 2013-2014. The Journal of infectious diseases 2016; 213 Suppl 3:S73–8.

16. Ouedraogo AL, de Vlas SJ, Nebie I, et al. Seasonal patterns of Plasmodium falciparum gametocyte prevalence and density in a rural population of Burkina Faso. Acta tropica 2008; 105:28–34.

17. Ouedraogo AL, Goncalves BP, Gneme A, et al. Dynamics of the Human Infectious Reservoir for Malaria Determined by Mosquito Feeding Assays and Ultrasensitive Malaria Diagnosis in Burkina Faso. The Journal of infectious diseases 2016; 213:90–9.

18. Derra K, Rouamba E, Kazienga A, et al. Profile: Nanoro Health and Demographic Surveillance System. Int J Epidemiol 2012; 41:1293–301.

19. Facebook Connectivity Lab and Center for International Earth Science Information Network - CIESIN - Columbia University. High Resolution Settlement Layer (HRSL). https://www.ciesin.columbia.edu/data/hrsl/ 2016.

20. Seattle - United States: Institute for Health Metrics and Evaluation (IHME). Global Burden of Disease Collaborative Network. Global Burden of Disease Study 2017 (GBD 2017) Population Estimates 1950-2017. http://ghdxhealthdataorg/record/ihme-data/gbd-2017-population-estimates-1950-2017 2015.

21. Held M, Karp RM. A Dynamic Programming Approach to Sequencing Problems. Journal of the Society for Industrial and Applied Mathematics 1962; 10:196–210.

22. Bennett K, Bradley P, Demiriz A. Constrained K-Means Clustering.

23. Zongo I OJ, Lal S, Cairns M, Scott S, Snell P, Moroso D, Milligan P. Coverage of SMC in Burkina Faso 2017. https://files.givewell.org/files/DWDA%202009/Malaria%20Consortium/SMC_in_Burkina_Faso_Coverage_surveys_2017.pdf 2018.

24. Blanford JI, Kumar S, Luo W, MacEachren AM. It’s a long, long walk: accessibility to hospitals, maternity and integrated health centers in Niger. Int J Health Geogr 2012; 11:24.

25. Makanga PT, Schuurman N, Sacoor C, et al. Seasonal variation in geographical access to maternal health services in regions of southern Mozambique. Int J Health Geogr 2017; 16:1.

26. Ba EH, Pitt C, Dial Y, et al. Implementation, coverage and equity of large-scale door-to-door delivery of Seasonal Malaria Chemoprevention (SMC) to children under 10 in Senegal. Sci Rep 2018; 8:5489.

27. Dougherty L, Abdulkarim M, Mikailu F, et al. From paper maps to digital maps: enhancing routine immunisation microplanning in Northern Nigeria. BMJ Glob Health 2019; 4:e001606.

28. Gidado SO, Ohuabunwo C, Nguku PM, et al. Outreach to underserved communities in northern Nigeria,2012-2013. The Journal of infectious diseases 2014; 210 Suppl 1:S118–24.

29. Kamadjeu R. Tracking the polio virus down the Congo River: a case study on the use of Google Earth in public health planning and mapping. Int J Health Geogr 2009; 8:4.

30. Kamanga A, Renn S, Pollard D, et al. Open-source satellite enumeration to map households: planning and targeting indoor residual spraying for malaria. Malaria journal 2015; 14:345.

31. Kelly GC, Seng CM, Donald W, et al. A spatial decision support system for guiding focal indoor residual interventions in a malaria elimination zone. Geospat Health 2011; 6:21–31.

32. Checchi F, Stewart BT, Palmer JJ, Grundy C. Validity and feasibility of a satellite imagery-based method for rapid estimation of displaced populations. Int J Health Geogr 2013; 12:4.

33. Oleribe OO, Momoh J, Uzochukwu BS, et al. Identifying Key Challenges Facing Healthcare Systems In Africa And Potential Solutions. Int J Gen Med 2019; 12:395–403.

34. Zon H, Pavlova M, Groot W. Regional health disparities in Burkina Faso during the period of health care decentralization. Results of a macro‐level analysis. Int J Health Plann Manag 2020; 35:939–59.

35. Guglielmi G. Africa declared free from wild polio - but vaccine-derived strains remain. Nature 2020.

36. The Alliance for Malaria Prevention (AMP). Considerations for distribution of insecticide-treated nets (ITNs) in COVID-19 affected countries. https://endmalaria.org/sites/default/files/Considerations%20for%20distribution%20of%20insecticide-treated%20nets%20%28ITNs%29%20in%20COVID-19%20affected%20countries.pdf 2020.

